# Clinical impact using low dose mycophenolate mofetil with tacrolimus on acute rejection, infectious diseases and non-infectious complications in renal transplant: A Single Hospital Experience in Mexico

**DOI:** 10.1101/2022.11.29.22282803

**Authors:** Jorge Andrade-Sierra, Hernesto Hernández-Reyes, Enrique Rojas-Campos, Ernesto G Cardona-Muñoz, José I Cerrillos-Gutiérrez, Eduardo González-Espinoza, Luis A Evangelista-Carrillo, Miguel Medina-Pérez, Basilio Jalomo-Martínez, Alejandra G Miranda-Díaz, Víctor M Martínez-Mejía, Benjamin Gómez-Navarro, Antonio de Jesús Andrade-Ortega, Juan J Nieves-Hernández, Claudia Mendoza-Cerpa

## Abstract

**Background:** The evidence supporting a starting dose of 2 g/day of mycophenolate mofetil (MMF) in combination with tacrolimus (TAC) in renal transplant (RT) is still limited but maintaining the dose of less than 2g could result in worse clinical outcomes in terms of acute rejection (AR).

**Research Question:** This study’s aim was to determine the association between AR, infectious and non-infectious complications after RT with the dose of 1.5g *vs*. 2g of MMF.

**Design:** A prospective cohort was performed with a 12-month follow-up in recipients of RT from living donors with low doses of MMF (1.5 g/day) or use of standard dosage (2 g/day). The association between adverse effects and complications and doses of MMF were examined in Cox proportional hazard models and survival free of AR, infectious diseases, and non-infectious complications was evaluated by the Kaplan-Meier test.

**Results:** At the end of the follow-up, the incidence of infectious diseases was 52% *vs*. 50% (*p*=0.71) and AR was 5% *vs*. 5% (*p*=0.86), respectively. Survival free of GI complications requiring medical attention was higher in the low dose group compared to the standard dose (88% *vs*. 45%, respectively; *p*<0.001).

**Discussion:** The use of 1.5 g/day of MMF is safe in Mexican population with RT from living donors without increasing the risk of AR, and it confers a reduction of adverse events.

## Background

Adverse reactions related to the immunosuppression used after renal transplantation (RT) could lead to infections and an increased mortality [1, 2], which has prompted attempts to use different treatment strategies in order to minimize immunosuppression and reduce these complications i.e. steroid reduction has been fully evaluated with controversial results. [3, 4]. In the standard immunosuppression scheme another option is mycophenolate mofetil (MMF) reduction. The recommended standard oral daily dose for adults is 2 g in RT and for the enteric-coated formulation, 720 mg of mycophenolate sodium (EC-MPS); is equivalent to 1 g of MMF.[5] However when the MMF is used in combination with tacrolimus (TAC) there is a lack in evidence supporting a starting dose of 2 g per day. Reducing the dose of MMF could be an option due to it gastrointestinal (GI) (diarrhea, abdominal pain, inflammatory colitis), hematological (neutropenia, anemia), and infectious adverse effects that have significant repercussions on quality of life and patient morbidity. [6-8] However, some of the retrospective studies using regimens with cyclosporine (CsA) and tacrolimus (TAC) suggest that MMF reduction and/or maintaining doses lower to 2g for a greater number of days, especially in the first year after RT, could result in worse clinical outcomes in terms of acute rejection (AR) [9-11]. A systematic review shows with weak evidence, that modifications (reduction or discontinuation) of MMF increase the rejection rate, and are more apparent in CsA-based regimens. [12] The use of TAC in the immunosuppressive drug regimen is first choice to recipients of RT[13]. Mycophenolic acid (MPA) is the active metabolite of MMF and TAC arises the exposure through lack of inhibiting the enterohepatic recirculation of the MPA glucuronide and in diabetic patients with delayed gastric emptying time this can contribute to a greater pharmacological exposure to MPA, [14-16]. Given the known pharmacokinetic and pharmacodynamic interactions whereby TAC increases MPA exposure and CsA does not, the concomitant use of TAC and MMF allows a theoretical reduction in MMF doses after RT with good results in terms of graft and patients survival during the first year after transplantation[17]; however, avoiding, suspending, or employing doses lower than 1g could increase the risk of AR.[18, 19] In our hospital, 97% of RT is from a living donor and the standard dose of 2g/day of MMF with TAC is used in the immediate post-RT period without taking into account the serum concentrations of MPA since this is impractical and economically unfeasible in our health care system considering the nonlinear absorption kinetics. Immunosuppression reduction is a clinical challenge, our experience in this field is based to steroid reduction [3, 4, 20], however clinical protocols in our setting allows to consider other pharmacological reductions i.e. MMF, there is an international evidence in MMF reduction with controversial[18, 19, 21-25] results, in Latin America there is not experience regarding this topic; Nevertheless, since the early post-transplant stages could considered the relatively low incidence of AR under the immunosuppressive regimens with TAC especially in low immunological risk patients clinicians find themselves having to reduce the doses of MMF, empirically, due to the presence of adverse effects. Thus, an actual tendency in our setting is to initiate, in the immediate post-transplant period, a dose of 1.5g/day. Consequently, faced with the lack of prospective studies related to the prescription of doses lower than the ‘standard’, we proposed the objective of determining the impact that the use of <2 g/day of MMF carries in infectious and non-infectious events and on AR in the first year of post-transplant follow-up.

## Methods

A prospective cohort study was performed in a single center (Transplant Division of the Specialties Hospital at the National Western Medical Center of the Mexican Institute of Social Security in Guadalajara, Jalisco, Mexico); from March 1st, 2017, to March 1st, 2018. All patients received a first renal transplant from living donor, and immunosuppressive maintenance regimen based on TAC, MMF and prednisone (PDN). Patients were divided into 2 groups according to the starting dose of MMF: low dose MMF (1.5 g/day) vs. standard dose MMF (2 g/day). All patients in our study were considered with low immunological risk, based on: a) negative results from cross-matching (flow cytometry), and b) pre-transplant negativity to anti-HLA antibodies to determine the absence of sensibilization. Treating nephrologists made the decision on induction type (thymoglobulin or basiliximab) according to clinical criteria; and the choice of MMF doses (low or standard doses) was determined by specific physician protocols, and this information was collected for the study. The exclusion criteria were as follows: multi organ recipients, re-transplants, deceased donor patients, pre-transplant medical or surgical gastrointestinal disorders, that could interfere with the absorption and distribution of immunosuppressants. The enzyme linked immunosorbent assay (ELISA) was used to evaluate the pre-transplant cytomegalovirus (CMV) status; the quantitative polymerase chain reaction (PCR) for CMV and decoy cell in urine; and immunohistochemistry for the sv40 antigen in renal tissue for the BK-virus was only used in case of clinical suspicion during follow-up. During the study period, all patients received prophylaxis with 900mg of valganciclovir for 3-6 months, and 160mg of trimethoprim and 800mg of sulfamethoxazole for 4 months.

### Information Collection and Analysis

The following data was collected: anthropometric characteristics, the presence of post-transplant infections (documented by clinical evaluation with systemic inflammatory response and fever, or confirmed by any microbiological method), hospitalizations, AR (documented with biopsy), glomerular filtration rate estimated (GFRe) with the study Modification of Diet in Renal Disease (MDRD) formula, and adverse events associated with MMF such as GI disorders (diarrhea, abdominal pain, nausea, vomiting, fullness, gastritis, anorexia, ulceration, hemorrhage), leukopenia (defined as leukocytes <4000 miles/uL), and/or anemia (defined as haemoglobin <11 g/dL). Our center has implemented as an ordinary practice, allograft biopsy protocols (gold standard) and for this study biopsies results were collected and the Banff’s 2017 Histopathology Classification was used. [26]

Immunosuppression induction was based on basiliximab 20 mg at 0- and 4-days post-transplant, or thymoglobulin at 1 mg/kg/day (accumulated dosage 3-4 mg/kg). Maintenance immunosuppression was based on: MMF 1.5g/day or MMF 2 g/day, TAC 0.10-0.12 mg/kg (to reach post-transplant target levels at days 1-30: 9-12ng/mL; and in 31-365 days: 8-10ng/mL), and starting prednisone (PDN) of 1 mg/kg/day with a reduction dose to achieve 0.1 mg/kg/day between the first and third post-transplant. Data are presented as mean ± standard deviation or median (percentiles 25-75%), and numbers and percentages where appropriate. Student *t* and Chi^2^ tests were used to compare groups. The Cox proportional hazards model was used to estimate the risk of development of infectious disease, AR, or non-infections complications in subjects with immunosuppressive regimens with low dose MMF *vs*. standard dose. Statistical analysis was performed with SPSS™ software, version 17 (SPSS, Inc., Chicago, IL). Survival free of AR, infectious diseases, and non-infectious complications was evaluated by the Kaplan-Meier test. All results with a value of *p*<0.05 were considered statistically significant.

#### Ethics approval

The present research complies with the Ethical Principles for Medical Research in Human Beings as stipulated in the Declaration of Helsinki 64th General Assembly, Fortaleza, Brazil, October 2013; in addition to adhering to the standards of good clinical practices. All procedures were performed according to national regulations as stipulated in the General Health Legal Guidelines for Health Care Research in Mexico, 2nd Title, in Ethical Aspects for Research in Human Beings, Chapter 1, and Article 17. The study was approved by the Ethics and Research Committee of the Specialties Hospital, National Western Medical Centre, Mexican Institute of Social Security (IMMS) in Mexico with registration number: R-2017-1301-108.

## Results

There were no significant differences between groups in terms of RT recipient’s gender, type and history of dialysis, co-morbidities, and HLA compatibility. Patients with low-dose MMF were younger and had a slightly lower body mass index (BMI) compared to those who received a standard dose. Table 1. Comparison of socio-demographic and transplant characteristics.TABLE1.docx

**Table 1.**
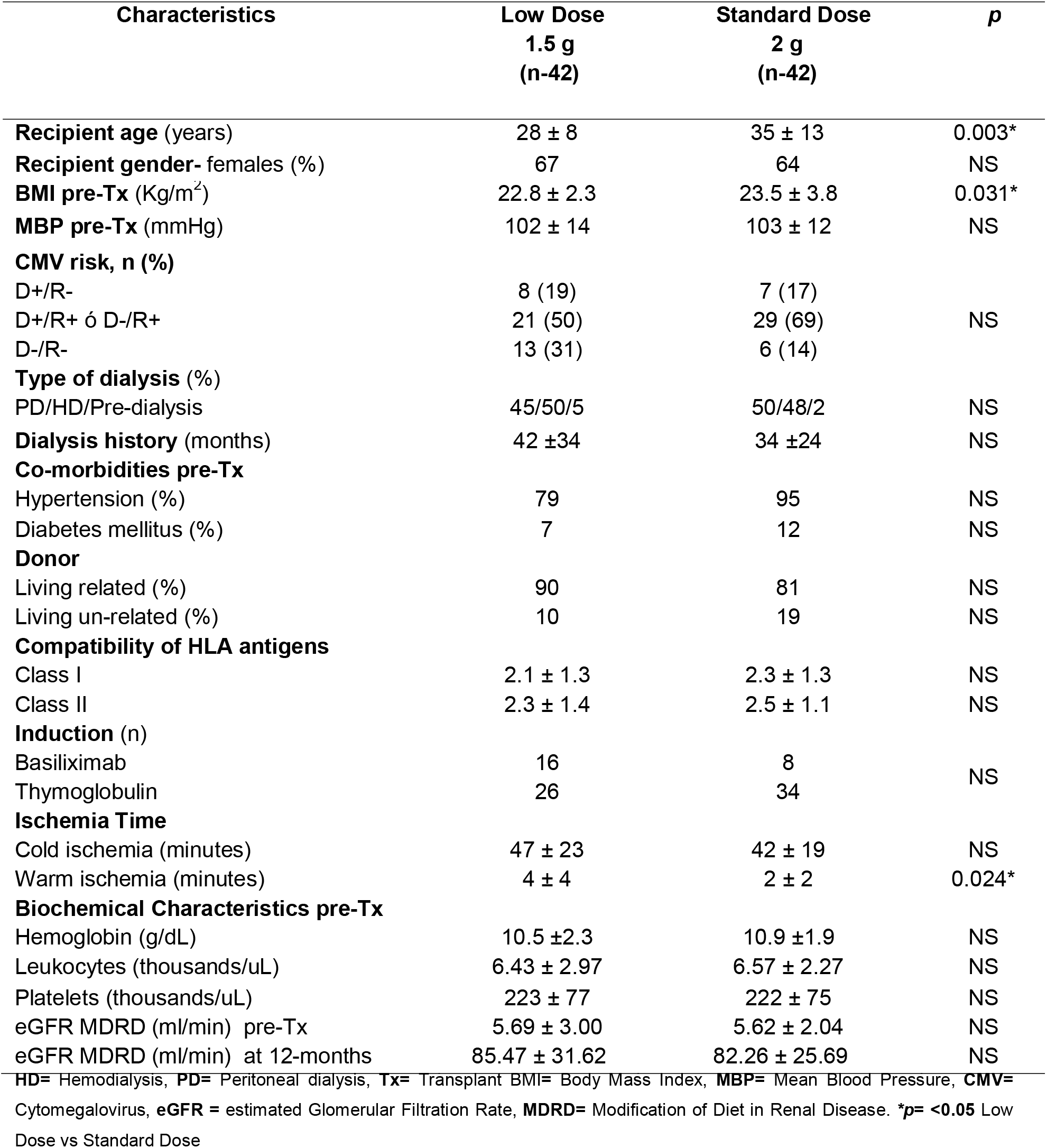
Comparison of socio-demographic and transplant characteristics

### Infectious complications

A survival analysis was performed to identify the infection-free time between both study groups, finding that within the first month of follow-up the greatest number of infectious events occurred in both groups. The incidence of infections was 52% for the low dose group and 50% for standard dose group, without statistical significance (Figure 1A). The urinary tract infections (UTI) predominated with 23 events and in 15 of the 23 events (65%) were secondary to Escherichia coli. The CMV infection occurred in 5 cases in the standard group and 1 case in the low doses group. In the UTI and CMV infections no statistical difference noted between the different MMF dose groups.

**Figure 1.**
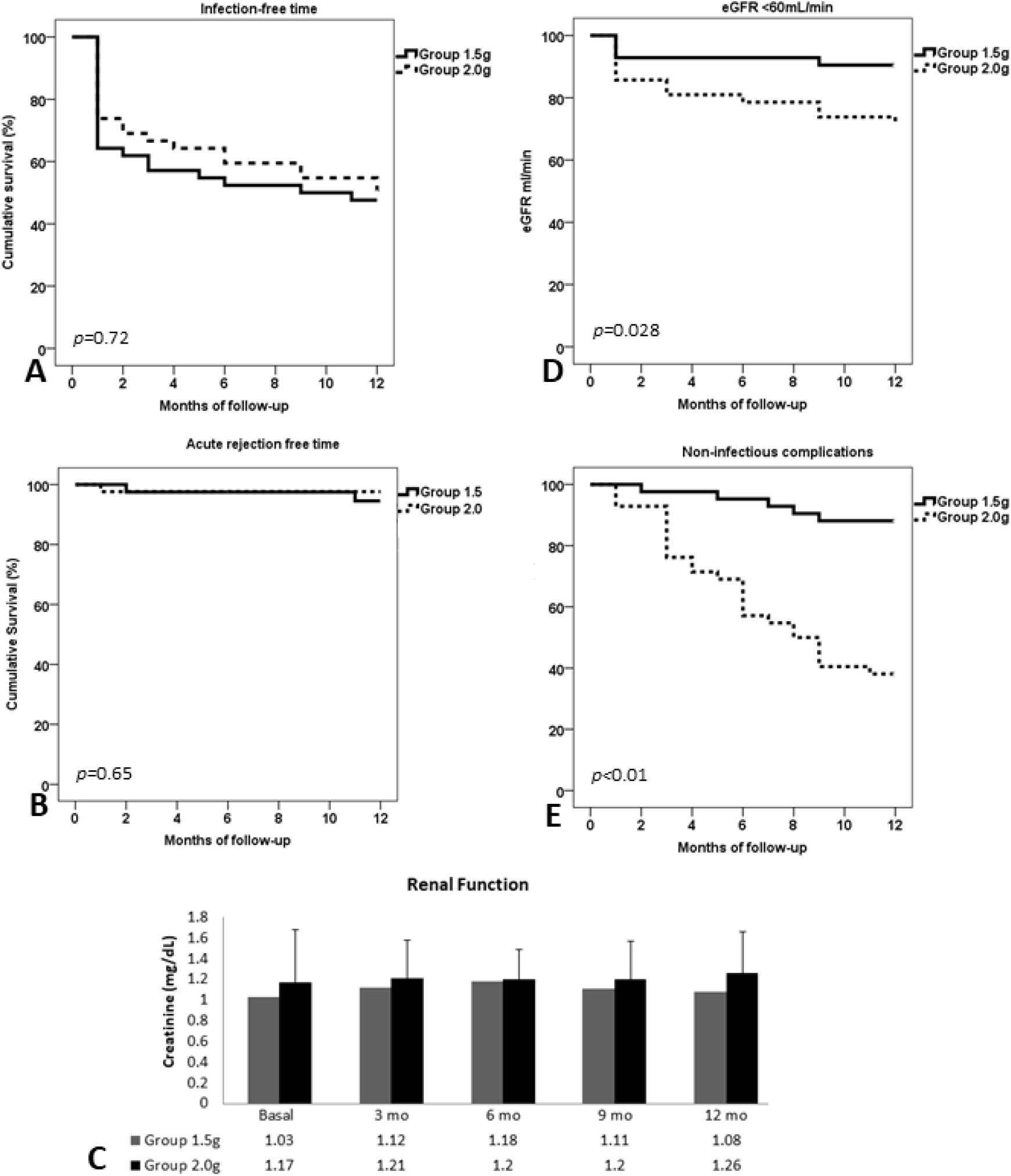
Infection-free time (1A), rejection-free time (1B), renal function at 12-month (1C), renal function establishing the eGFR with a cut-off point at <60 mL/min/m^2^ (1D) and non-infectious complications (1E). Comparations between both study groups.

### Immunological complications

Likewise, with the aim of assessing the rejection-free time during the 12-month follow-up, a survival analysis was performed, the incidence was similar with 2 events recorded during the follow-up in both study groups (4.7%, p=NS), and achieving an AR-free survival of 95% at the 12-month follow-up (p=0.28) (Figure 1B). Evaluation of the graft function at the 12-month follow-up was estimated by the measurement of serum creatinine. Patients treated with low dose MMF showed no difference in renal function at 12-month (Figure 1C), however the standard dose group seems to have lower eGFR, therefore, a secondary analysis was carried out with the objective of identifying the outcome of renal function in both study groups and establishing the eGFR with a cut-off point at <60 mL/min/m^2^. The results of this sub-analysis showed that for the low dose group survival [free of this event] occurred in up to 90% of the cases *vs*. 71% of the survival free of the event in the standard dose group at the 12-month follow-up (*p*=0.028) (Figure 1D). Figure 1..tif

### Hematologic complications

The time free of leukopenia at the 12-month follow-up was found in 78% and 69% (*p*=0.34) in the group with 1.5g *vs*. 2 g, respectively. Likewise, the time free of anemia was found in 64% (MMF 1.5g/day) and in 45% (MMF 2g/day) (*p*=0.08). These hematologic abnormalities, such as anemia, can occur with MMF, but it is possible with any myelosuppressive therapies, like with the use of antithymocyte globulin or valgancyclovir. We do not find some association with these drugs. It is possible that the dose reductions or modifications of MMF were performed to the adverse effect of MMF (information not analyzed).

### Gastrointestinal (GI) complications

The time free of GI complications (fullness, nausea, gastritis, anorexia, abdominal pain, diarrhea) presented in 88% of the group treated with MMF 1.5g/d and 45% in the group treated with MMF 2 g/d (*p*=0.001)(Figure 1E). Figure 1..tif

The figure 1 shows the comparations between both study groups with respect to infection-free time (1A), rejection-free time (1B), renal function at 12-month (1C), renal function establishing the eGFR with a cut-off point at <60 mL/min/m^2^ (1D) and non-infectious complications (1E). Figure 1..tif

Analysis of risk of the predictive variables for eGFR <60mL/min showed the dose of 1.5g/d (RR=0.29, CI 95% 0.09-0.91, *p*=0.04) and renal function on discharge (RR=0.96, CI 95% 0.94-0.98, *p*=0.001) as a protective (Table 2). TABLE2.docx

**Table 2.** Risk factors associated with eGFR < 60mL/min at 12-month of follow-up

| **Factor** | **RR** | **95% CI** | ***p*** |
| --- | --- | --- | --- |
| 1.5 g **MMF** | 0.29 | 0.09-0.91 | 0.04 |
| **Age** (donor) | 1.05 | 1.01-1.09 | 0.02 |
| **MDRD** at discharge | 0.96 | 0.94-0.98 | 0.001 |
| Chi<sup>2</sup>= 19.78; *p*<0.0001 |  |  |  |

Adjusted for number of antigens, age (receptor), dialysis time, ischemia time, body mass index (BMI), induction therapy (Basiliximab/Thymoglobulin)

Analysis of the predictive data for GI complications showed that at a dose of 1.5 g/d (RR=0.11, CI 95% 0.040-0.029, *p*=0.0001) and type of donor (live, related) (RR=0.29, CI 95% 0.11-0.77, *p*=0.013) were protective factors (Table 3).TABLE3.docx During follow-up, the patients with initial dose of MMF 2.0g/day required more dosage adjustments associated with non-infectious complications (Gastrointestinal).

**Table 3.**
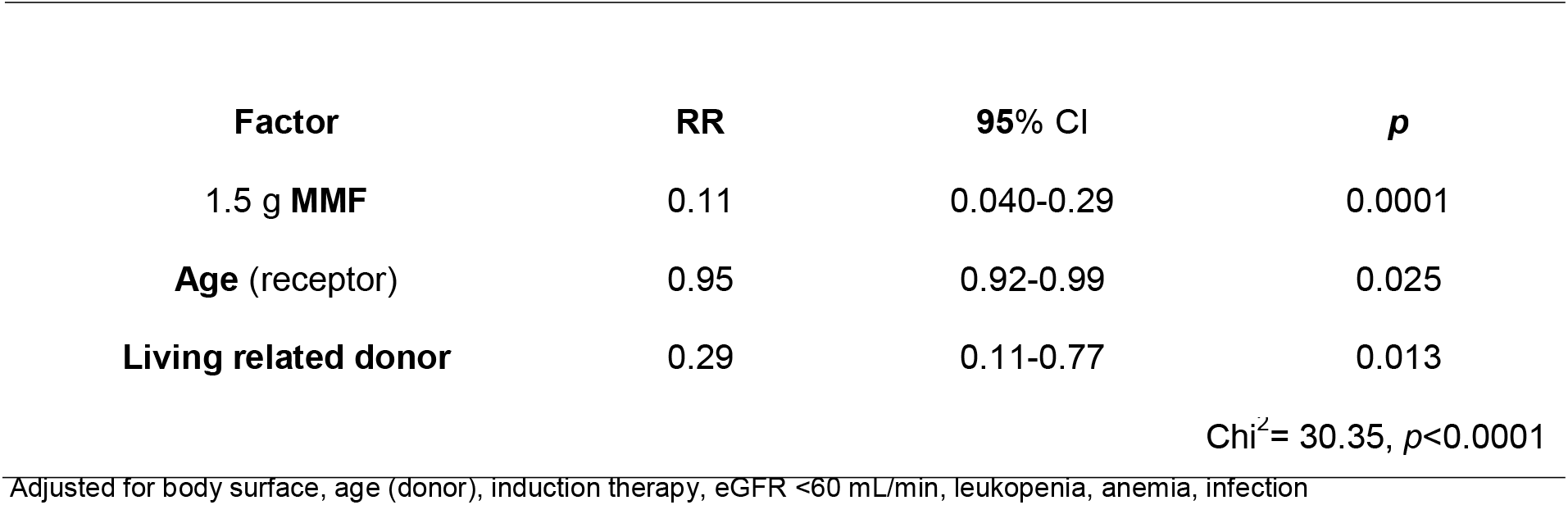
Risk factors associated with gastrointestinal complications

| **Factor** | **RR** | **95% CI** | ***p*** |
| --- | --- | --- | --- |
| 1.5 g **MMF** | 0.11 | 0.040-0.29 | 0.0001 |
| **Age** (receptor) | 0.95 | 0.92-0.99 | 0.025 |
| **Living related donor** | 0.29 | 0.11-0.77 | 0.013 |
| Chi<sup>2</sup>= 30.35, *p*<0.0001 |  |  |  |

Adjusted for body surface, age (donor), induction therapy, eGFR <60 mL/min, leukopenia, anemia, infection

## Discussion

Immunosuppression reduction is a complex field in transplant; our setting has experience in this field; according to our previous findings steroid reduction has not associated to AR increment [3, 20], however is related to antibodies development.[4] To the best of our knowledge this is the first study of MMF reduction in Latin-America. The infectious events (UTI, CMV, BK), neoplasms, GI and hematological toxicity (leukopenia), are associated with the use of immunosuppression based in MMF in retrospective analyses [8, 10, 21, 27]; therefore, the minimization of MMF seems to be a therapeutic option with the aim of reducing these complications that impact quality of life and morbidity-mortality in RT recipients. [6, 7]

The proportions of UTI infection tended to be higher in the low dose group in our study, while CMV infection was higher in the 2g/d group, but neither demonstrated significant differences between the groups. Most cases were pyelonephritis secondary to Escherichia Coli and occurred during both the perioperative period and the late postoperative period and responded to the appropriate antibiotic therapy without affecting graft survival. In the cases of CMV, the infection provoked significant reductions of the doses compared to the low dose group.

In retrospective analysis by Hosohata *et al*. [8], of the different adverse events that were recorded in adult RT, 45.8% were associated with MMF and the principal infectious events were CMV, UTI, and BK with statistical significance. There are many reasons for MMF reduction or withdrawal. Park *et al*. [18], in a retrospective analysis with induction and maintenance immunosuppression similar to our study, found that the common causes of discontinuation of MMF were infections (70.7%) but when they analyzed the different groups (continuation or withdrawal of MMF) in the long term, they no found differences in the incidence of CMV and BK. In the present cohort, we found no statistical significance with UTI and CMV between the groups, and BK nephropathy was not demonstrated through decoy cells in any histopathological result; although, the disadvantage of our study was lack of measurement of the viral load for BK. In our study, survival free of infectious illnesses in general was not statistically different between the two different doses employed (52% in the group with 1.5 g/day *vs*. 50% in the 2.0 g/day group).

Our results concur with those of Doria *et al*. [28] who demonstrated that the initial doses of <2g of MMF subpopulations did not compromise the safety of the renal graft at 12 months of follow-up, with a similar percentage of discontinuation of study drug between EC-MPS and MMF treated patients by month 12 with an immunosuppressant regimen similar to ours based in TAC; establishing that the dose of 1.5g, from the immediate post-transplant period, is an adequate therapeutic strategy. The concomitant use of TAC and MMF favors the exposure to MPA compared with CsA as a consequence of different mechanisms (lack of inhibition of enterohepatic MPA, drug-drug interaction with MPA metabolism) [14-16], allowing for reducing doses of MMF in TAC-treated patients. Ji SM *et al*.[19] with regimens based in TAC, PDN, and MMF, showed greater risk of AR with Cd4 deposits when using a dose of <1.5g. Similarly, the withdrawal of MMF is a risk for AR. Park *et al*.[18], showed that the proportion of AR was significantly higher when the MMF is taken away and is associated as an independent risk factor for graft failure with death-censored graft survival significantly lower compared to the group where MMF is continued. On the other hand, Vanhove T et al[21]; showed MMF dose reductions of ≥50% independently increased the risk of AR but did not compromise graft survival. Similar results at 2 years were reported by Kocak H et al[22] using a dose of 1 gr with better survival and patient survival rates than higher MMF dose.

In the same way the study THOMAS demonstrated that MMF could be even discontinued from a TAC based triple regimen 3 months after transplantation in low immunological risk patients without negative impact on AR rate, graft and patient survival, and renal function. [23, 24]

In patients from the lower dose group was not necessary a greater reduction, despite the fact that those with standard dose experience a significant reduction in MMF doses however none were lower than 1.5g in either group. However, in contrast to findings from a retrospective analysis reported by Dave V *et al*., [25] we did not find that an initial low dose of MMF was associated with an increase in the incidence of AR, although the population studied in this analysis was different (45% deceased donor recipients, 17% with panel reactive antibodies (PRA) higher 20% and ABO blood group incompatible in 22%. We only evaluate survival-free of AR during follow-up (12 months), suggesting that the initial dose of 1.5g of MMF does not lead to increased risk of AR a short-term in our population. The MMF dose of 1.5g does not appear to affect graft survival, above all with the concomitant use of TAC. [25, 29, 30] The present study used the immunosuppressant maintenance regimen of TAC, PDN, and MMF, and we did not lose any grafts in both groups; however given the methodological limitation (this is a Cohort study), we did not consider this as final response. It is necessary to evaluate this finding in a clinical trial; however, there is not any study in Latin America that could provide the evidence we found in this Cohort. Our results are similar to other populations that included both living and deceased donor recipients with low immunological risk[25, 28]. In fact in the Doria’s study the proportion of African Americans and patients with PRA <30% was similar in the cohorts receiving <2 g or 2 g, as was the proportion of living donors.[28] Similarly Dave et al[25]; despite the fact that a dose of 1.5 g (adjusted for the immunological risk of the transplant) was associated with AR no showed difference in graft survival or renal function. During follow-up our cohort did not lose any grafts with reduction MMF dose up to 1.5 g/day, and sustain an eGFR >60mL/min for a longer period of time.

The Cox hazard proportional risk analysis showed that reduced MMF dose (1.5 g/day), was able to sustain an eGFR >60mL/min for a longer period of time. On the other hand, there was a slightly difference in baseline BMI between two study cohorts, and this difference may influence physicians in the decision to use low doses of MMF; however, the standard dose group markedly reduced MMF dose throughout the year of follow-up while those who received low doses did not change during that time. Body weight could not be a critical variable in clinical practice, due to the great inter-patient variability of pharmacokinetics parameters, especially when is co-administered with TAC. There is evidence regarding that is possible even 10-fold variation of MPA exposure in patients receiving the same MMF dose. [31]

The possibility to obtain better MPA levels and less variability in the adjustment of the MMF dose is also achieved with the concomitant use of TAC in immunosuppressant regimens, since TAC favors greater plasma exposure of MPA.[15, 16] Walker *et al*. [32], showed a high incidence of AR in failing to achieve adequate TAC concentrations, and suggest that a high target TAC concentration within the first week after renal transplantation in order to achieve the early minimization of MMF in combination with TAC. In our study, the proportion of patients treated with TAC was not modified, and the TAC levels were sustained during follow-up.

The GI toxicity is frequent and dependent on MMF dose. [33] Tierce *et al*.,[11] in a retrospective analysis, showed 49.7% of GI complications in the first six months requiring 39% dose adjustment or discontinuation, and with higher AR compared to patients without GI complications. In our study, the group treated with 1.5g of MMF had lower weight and body surface area; therefore, we assumed that the clinics took that data into account when dosing the medication. However, the group that received 2.0g of MMF, during, and at the end of follow-up, had significant reductions of the dose due to adverse effects. Contrary to the observational study with different doses (< 2g, 2g, and >2g) analysis of the association revealed significantly higher GI adverse events requiring discontinuation in 10.1% of MMF-treated patients by the twelfth month. [28] In our study, survival free of GI complications was higher in the low dose MMF group while with the standard dose these were the main cause for seeking medical attention and the principal motive for adjusting the medication dose. Bunnapradist *et al*., associated the female gender, the presence of diabetes, and concomitant use of MMF and TAC, with non-infectious diarrhea [6]. In our analysis of prediction of risk, we found at dose of 1.5 g/day MMF and the type of donor (live related) were protective for GI complications.

Anemia and leukopenia, with or without neutropenia, are frequent in the first post-transplant months.[6, 7, 27] In a retrospective study, Knoll *et al*. [10], found leukopenia (55.1%) as the main cause of reducing the dose of MMF followed by GI complications (22%). In our results, the time free of leukopenia at 12 months of follow-up was lower in the standard group, and was a reason to reduce the dose. However, differences in the hematologic complications (anemia and leukopenia) were not statistically significant between the different doses of MMF.

Recently, Michielsen *et al*.,[34] demonstrated that independently of the immunosuppression initiated from the immediate post-transplant period, the long-term survival of the grafts is excellent; even with less potent immunosuppressant regimens. In this sense, we consider that reduction of the MMF dose with schemes based on TAC and PDN could be feasible; although prospective studies would be necessary to evaluate long-term survival in our population.

Asians tend to exhibit a higher MPA exposure in the early post-transplant period suggesting the need for another MMF dose in this population. [35] In the Hispanic population there is little information reported but the exposure appears to be high too [36]. We considered that body weight might be an important factor contributing to ethnic difference, without disregarding the role of other variables.

### Limitations

We would like to acknowledge the limitations of our study; on one hand its only one center character which, although it can favor consistency in MMF dosing in our country and Latin populations, limits the extrapolation of results to other populations. This does not reduce the validity, but its interpretation requires the consideration of local conditions and circumstances. Despite our findings reinforcing the concept to use a lower dose of MMF safely in combination with TAC and PDN in our population, the lack of random assignment of the initial MMF dose only permits us to speculate on potential reasons that can explain observed differences between the different doses employed. The results provide information about a topic that still needs information in different populations; however, the results should be validated in our center through randomized clinical trials. On the other hand, the absence of follow-up of the therapeutic levels of MPA in our country due to economical and logistical limitations is an irrefutable reality. The small sample size and short follow-up of the study could be limitations; however the follow-up in the short term (first post-transplant year) evaluate the time where a greater risk of AR and/or adverse effects of MMF exist and finally; the possible causes of noncompliance were not controlled.

## Conclusions

Clinical management using the dose of 1.5g of MMF from the immediate post-transplant period in living-donor RT recipients, with immunosuppression based in TAC and PDN is safe, without increasing the risk of AR, and with a reduction in the GI adverse events that require hospitalization. These findings need to be validated with clinical trials, but we consider that the dose reduction could be safe if there is strict vigilance in watching for potential adverse effects through frequent clinical, biochemical, and histopathological assessments.

## Data Availability

Data availability
Most clinical data are restricted to protect information against misuse. The data may be shared by the Mexican Social Security Institute (IMSS) after approval from the Ethics Committee of the institution. The data that support the findings of this study are available from the corresponding author upon reasonable request.

## Acknowledgments

Not applicable.

## Declaration of Conflicting Interests

The authors declared no potential conflicts of interest with respect to the research, authorship, and/or publication of this article

## Funding

The study did not receive any private or governmental funding.

## Authors’ contributions

a. ASJ, HRH, RCE : *Participated in the conception and design* of the study.
b. ASJ, HRH, RCE, CMEG, CGJI and *BGN: Analyzed and interpreted the data*.
c. *GEE, ECLA, MPM, MDAG, MMV, AJAO, NEJJ and MCC: Participated in the drafting the article and critical review for important intellectual content*.
d. *Final approval of the version to be published*: All the team

## Notes

### Competing Interest Statement

The authors have declared no competing interest.

### Author Declarations

The study was approved by the Ethics and Research Committee of the Specialties Hospital, National Western Medical Centre, Mexican Institute of Social Security (IMMS) in Mexico with registration number: R-2017-1301-108.

